# National impact of Score MAMÁ on obstetric complications in Ecuador, 2012–2024: interrupted time series analysis

**DOI:** 10.64898/2026.10.20.26366065

**Authors:** Andrés Sebastián Herdoíza Almeida, Luis Gustavo Paredes, Marcelo Alberto Guerrero Salinas, Alex Wladimir Guayta Valladares, Andrés Alejandro Trujillo Solórzano, Enrique Santiago Lanas Mendoza

## Abstract

**Objectives:** To evaluate the impact of Score MAMÁ, Ecuador’s national obstetric early-warning system, on rates of severe obstetric complications (hypertensive, hemorrhagic, septic), comparing the 2015 pilot phase (Zones 4/9) with the 2016 national rollout.

**Methods:** Quasi-experimental ecological interrupted time series study using national administrative data from Ecuador (2012–2024). The denominator came from the live-birth registry (ENV-INEC; n = 3 581 602); the numerator came from hospital discharge records (EDG-INEC), classified by ICD-10 code as Blue Code (O14–O15), Red Code (O44–O46/O67/O72), or Yellow Code (O85–O86/O41.1), aggregated monthly across Ecuador’s 9 SENPLADES zones. Negative binomial models with a live-birth offset and zone-nested first-order autoregressive (AR1) correlation estimated incidence rate ratios (IRR). A secondary national-annual analysis modeled maternal mortality by code.

**Results:** We identified 200 075 discharges for an obstetric complication (Blue = 125 848; Red = 55 049; Yellow = 19 178). Blue Code increased in the pilot phase (IRR 1.27; 95% CI 1.03–1.57) and nationally (IRR 1.20; 95% CI 1.09–1.33); Red and Yellow Code showed no change. A national mortality analysis (817 deaths) found no significant change for any code (IRR 0.76–1.62; CI spanned 1).

**Conclusions:** Score MAMÁ was associated with a selective increase in recorded hypertensive complications, with no change in hemorrhage, sepsis, or maternal mortality — a pattern more consistent with improved detection of preeclampsia-eclampsia than a true change in complication incidence.

## Introduction

Maternal mortality remains one of the most sensitive indicators of health inequity worldwide. A recent World Health Organization systematic analysis of causes of maternal death for 2009–2020 identified hemorrhage as the most frequent cause (27%), followed by indirect obstetric causes (23%) and hypertensive disorders of pregnancy (16%), with the proportion of deaths from hypertensive disorders notably higher in Latin America and the Caribbean than in other regions (1). While mortality from hemorrhage has declined steadily over recent decades, progress against hypertensive disorders has been far more limited, leading to projections that these disorders will become the leading cause of maternal mortality worldwide if current trends persist (2).

Against this backdrop, obstetric early-warning systems have been proposed as a low-cost strategy to standardize the clinical recognition of maternal deterioration, building on the United Kingdom’s confidential enquiries into maternal deaths and scales such as the Modified Early Obstetric Warning System and the ICNARC Obstetric Early Warning Score (3,4). Validation studies in comparable regional settings, such as one in Colombia using the ICNARC-OEWS, showed good discriminative performance for predicting maternal death among critically ill patients (AUC 0.84–0.87) (5). More recent systematic reviews are more cautious: this remains a promising tool requiring further evidence and standardization (6), and the WHO Global Maternal Sepsis Study found that no scale alone achieved sufficient diagnostic accuracy for sepsis, though obstetric-specific scales outperformed generic ones (7) — suggesting these systems’ performance may not be uniform across complication types.

Within this context, Ecuador’s Ministry of Public Health designed Score MAMÁ, a national obstetric early-warning system organized around three “codes” — Blue (hypertensive disorders), Red (hemorrhage), and Yellow (sepsis) — each with standardized response protocols. The system was formally approved in 2017, piloted in SENPLADES Zones 4 and 9 in August 2015, and scaled up nationally in January 2016 (8); it was updated in 2025 as a further effort to reduce maternal morbidity and mortality through more standardized clinical processes (17). This staggered, non-experimental rollout constitutes a natural quasi-experimental opportunity to evaluate population-level impact. Beyond its clinical protocols, by embedding a common blue/red/yellow nomenclature into routine documentation, Score MAMÁ plausibly also standardized how these events are coded and reported administratively — relevant to interpreting any observed change in discharge rates.

To our knowledge, no nationwide quantitative evaluation of the impact of Score MAMÁ on rates of severe obstetric complications has been published. The present study aims to fill that gap through an interrupted time series analysis using national administrative records of hospital discharges and live births between 2012 and 2024. We hypothesized that if Score MAMÁ primarily improves clinical detection rather than primary prevention, its effect should be larger for the code that depends on active, systematic screening (Blue) than for codes with acute clinical presentations that are unlikely to go unnoticed (Red and Yellow).

## Methods

### Design and setting

We conducted a quasi-experimental ecological interrupted time series study, with the zonal level (Ecuador’s 9 SENPLADES planning zones) as the unit of analysis and monthly periodicity, from January 2012 through December 2024 (156 months). The intervention of interest was Score MAMÁ, with two temporal milestones: the pilot phase, beginning in August 2015 in Zones 4 and 9 (pilot group), and the national rollout, beginning in January 2016 for the rest of the country (control group during the pilot phase).

### Data sources and case definition

The denominator was constructed from Ecuador’s Live Birth Statistics registry (ENV-INEC) for 2012–2024, after a data-cleaning process, harmonization of cantonal geographic codes (4-digit DPA code), and exclusion of records with sentinel or unresolved codes (final n = 3 581 602 live births across 219 cantons).

The numerator was constructed from Hospital Discharge records (EDG-INEC) for the same period, harmonizing 13 annual files with heterogeneous formats and coding levels. Discharges with a principal diagnosis in women were classified by ICD-10 code into one of three obstetric codes, following the Score MAMÁ protocol (8,17): Blue Code (hypertensive disorders, O14–O15, across their full severity spectrum), Red Code (obstetric hemorrhage, O44–O46, O67, O72), and Yellow Code (pregnancy-related sepsis, O85, O86, O41.1). After excluding cases with an unresolvable cantonal code (0.14%), we identified 200 075 discharges for a Score MAMÁ-related obstetric diagnosis (Blue = 125 848; Red = 55 049; Yellow = 19 178), aggregated monthly by canton and consolidated into the 9 SENPLADES zones using a validated geographic crosswalk. The numerator and denominator are not strictly commensurable: the numerator counts qualifying discharges, normalized to live births as the best available population-at-risk proxy, not a direct incidence measure — a single pregnancy can generate more than one qualifying discharge (e.g., an antepartum hypertension admission followed by a separate delivery admission), and the count includes discharges not linked to a live birth (e.g., stillbirths). We therefore refer to discharge or diagnosis rates rather than complication incidence in the strict epidemiological sense.

### Statistical analysis

For each obstetric code, we constructed a complete zone-month panel (including months with zero cases; 1 404 zone-month observations per code) and modeled the monthly count of discharges using negative binomial regression (type 2, constant-dispersion parameterization) with an offset for the log of monthly live births in the corresponding zone, following the segmented-regression logic for interrupted time series described by Wagner et al. (9) and Taljaard et al. (10). For zone z and month t (t = 1, …, 156; January 2012–December 2024), the model was specified as:

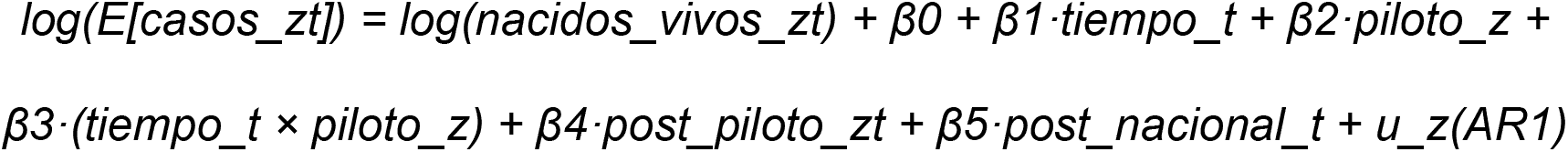

where piloto_z equals 1 for the pilot zones (Z4/Z9) and 0 otherwise (time-invariant); post_piloto_zt equals 1 for pilot-zone observations from August 2015 onward and stays 1 for those zones after the national rollout, since the indicator is not switched off; post_nacional_t equals 1 for all zones from January 2016 onward; and u_z(AR1) is a zone-nested first-order autoregressive random effect over the monthly time index. Because post_piloto and post_nacional are both coded 1 for pilot zones after January 2016, they are additive, non-mutually-exclusive level shifts: exp(β4), the “Pilot (Z4/Z9)” IRR in Table 1, is the incremental shift specific to pilot zones from August 2015, over and above the subsequent national effect; exp(β5), the “National” IRR, is the shift applying countrywide from January 2016. The model has no separate post-intervention slope-change terms, only step-level shifts; exp(β3), the constant pilot-vs-rest trend difference across the full period, was examined as a pre-intervention parallel-trends check rather than reported as an effect estimate.

**Table 1.** Incidence rate ratios (IRR) for the association between Score MAMÁ implementation phases and hospital discharge rates.

| Code | Intervention phase | IRR | 95% CI | p value |
| --- | --- | --- | --- | --- |
| Blue<br>(hypertensive) | Pilot (Z4/Z9) | 1.27 | 1.03 – 1.57 | 0.025 |
|  | National | 1.20 | 1.09 – 1.33 | <0.001 |
| Red<br>(hemorrhagic) | Pilot (Z4/Z9) | 1.03 | 0.77 – 1.39 | 0.826 |
|  | National | 0.93 | 0.81 – 1.07 | 0.300 |
| Yellow (septic) | Pilot (Z4/Z9) | 0.96 | 0.63 – 1.46 | 0.856 |
|  | National | 0.98 | 0.80 – 1.20 | 0.842 |

*Note. IRR: incidence rate ratio; CI: confidence interval; AR1: first-order autoregressive. Pilot (Z4/Z9): SENPLADES Zones 4 and 9, piloted from August 2015; National: nationwide rollout from January 2016. Blue Code: hypertensive disorders (ICD-10 O14–O15); Red Code: obstetric hemorrhage (O44–O46, O67, O72); Yellow Code: pregnancy-related sepsis (O85–O86, O41.1).*

Initial models without a correlation structure showed severe month-to-month residual autocorrelation within each zone (Durbin-Watson predominantly 0.13–1.2), so the final model incorporated the zone-nested AR1 term shown above, fitted by maximum likelihood in glmmTMB (negative binomial type 2) in R. This primary specification excludes calendar-month fixed effects; a seasonally adjusted variant (adding an 11-category month factor) was fitted as a sensitivity check. Model adequacy was assessed with simulated residuals (DHARMa package), evaluating overdispersion and autocorrelation. Coefficients were exponentiated to obtain IRR with 95% CI.

### Sensitivity analyses

We conducted five further sensitivity analyses: a pre-intervention (before August 2015) parallel-trends check; ACF/PACF residual diagnostics justifying the AR1(1) structure; the seasonally adjusted variant above, plus an alternative linear-dispersion (negative binomial type 1) specification; two COVID-19 checks (a 2020–2021 pandemic-year indicator, and separately, excluding 2020–2021); and a negative control outcome not targeted by Score MAMá’s protocols (ICD-10 O24, diabetes complicating pregnancy), modeled identically. We also projected national monthly observed, fitted, and counterfactual (intervention terms fixed at 0) Blue Code discharge rates per 10 000 live births, to express its effect on an absolute scale.

### Mortality analysis

Score MAMÁ’s own stated objectives explicitly include preventing maternal death, so whether the intervention is associated with a detectable change in mortality is a question this evaluation should address directly rather than defer. We therefore performed a secondary, exploratory analysis of national maternal mortality using Ecuador’s General Mortality Statistics registry (EDG-INEC) for 2012–2024 (1 044 128 deaths of women of all causes and ages). Deaths were classified by the 4-character ICD-10 underlying-cause code using the same code-specific definitions used for the complications numerator (Blue: O14–O15; Red: O44–O46, O67, O72; Yellow: O85–O86, O41.1), restricted to female decedents. This classification was validated against INEC’s own official maternal-death flag, available for 2012–2014 only; agreement was exact for 2012 (205/205 true maternal deaths and 63 306/63 306 true non-maternal deaths correctly classified).

Unlike the complications analysis, the mortality analysis could not be conducted at the zonal level: the canton-of-residence field needed to distinguish the pilot zones (Z4/Z9) and the Zone 8 cantons (Guayaquil, Durán, Samborondón) from the rest of their provinces is present only for 2012–2014; from 2015 onward only canton of death, canton of civil registration, and province of residence are retained. Combined with the rarity of maternal death (national annual counts of 3–59 per code), this precluded a reliable zone-month model. We instead fit a national, annual-resolution segmented regression (13 observations per code, 2012–2024), following the same segmented-regression logic (9,10) but with a single breakpoint at the January 2016 rollout; the August 2015 pilot phase, affecting 2 of 24 provinces for five months, could not be isolated at this resolution. The denominator was the same live-birth registry, aggregated nationally by year. Poisson regression with a live-birth offset was the base model per code; negative binomial was substituted when the Pearson dispersion statistic exceeded 1.5. We additionally fit an aggregate model pooling all three codes, reported as “Pooled (Blue+Red+Yellow)”.

### Ethical considerations

This study used exclusively secondary, aggregated, and anonymized administrative data, with no individual patient identification. The study protocol was reviewed by the Research Ethics Committee on Human Subjects (CEISH, Comité de Ética de Investigación en Seres Humanos) of the International University of Ecuador (UIDE), the institutional ethics committee of record for this study, which determined that the exclusive use of secondary, aggregated, de-identified administrative data, with no direct patient contact, qualified the study for exemption from full committee review.

## Results

Between January 2012 and December 2024, 3 581 602 live births and 200 075 hospital discharges for one of the three obstetric codes under study were recorded, distributed across Blue Code (n = 125 848; 62.9%), Red Code (n = 55 049; 27.5%), and Yellow Code (n = 19 178; 9.6%). The zonal analysis included 9 planning zones and up to 156 months per zone (1 404 zone-month observations per code).

All three primary (non-seasonal) models showed very high month-to-month within-zone correlation (0.96 Blue, 0.97 Red, 0.98 Yellow), confirming that correcting for autocorrelation was indispensable. DHARMa diagnostics showed no residual overdispersion for Blue Code (p = 0.12), whereas Red and Yellow Code retained significant residual overdispersion (p < 0.001 and p = 0.008). Because overdispersion tends to shrink standard errors and favor false positives, its persistence for Red and Yellow is an unresolved model-fit limitation, not evidence reinforcing the null finding; it is addressed with additional specifications in a dedicated sensitivity analysis.

Table 1 and Figure 1 summarize the estimated incidence rate ratios for the pilot phase and the national phase for each obstetric code. Blue Code showed a statistically significant increase in both phases: 27% during the pilot phase in Zones 4/9 (IRR 1.27; 95% CI 1.03–1.57; p = 0.025) and 20% following the national rollout (IRR 1.20; 95% CI 1.09–1.33; p < 0.001). By contrast, neither Red Code nor Yellow Code showed a significant change in either phase, with confidence intervals that broadly spanned the null value.

**Figure 1.**
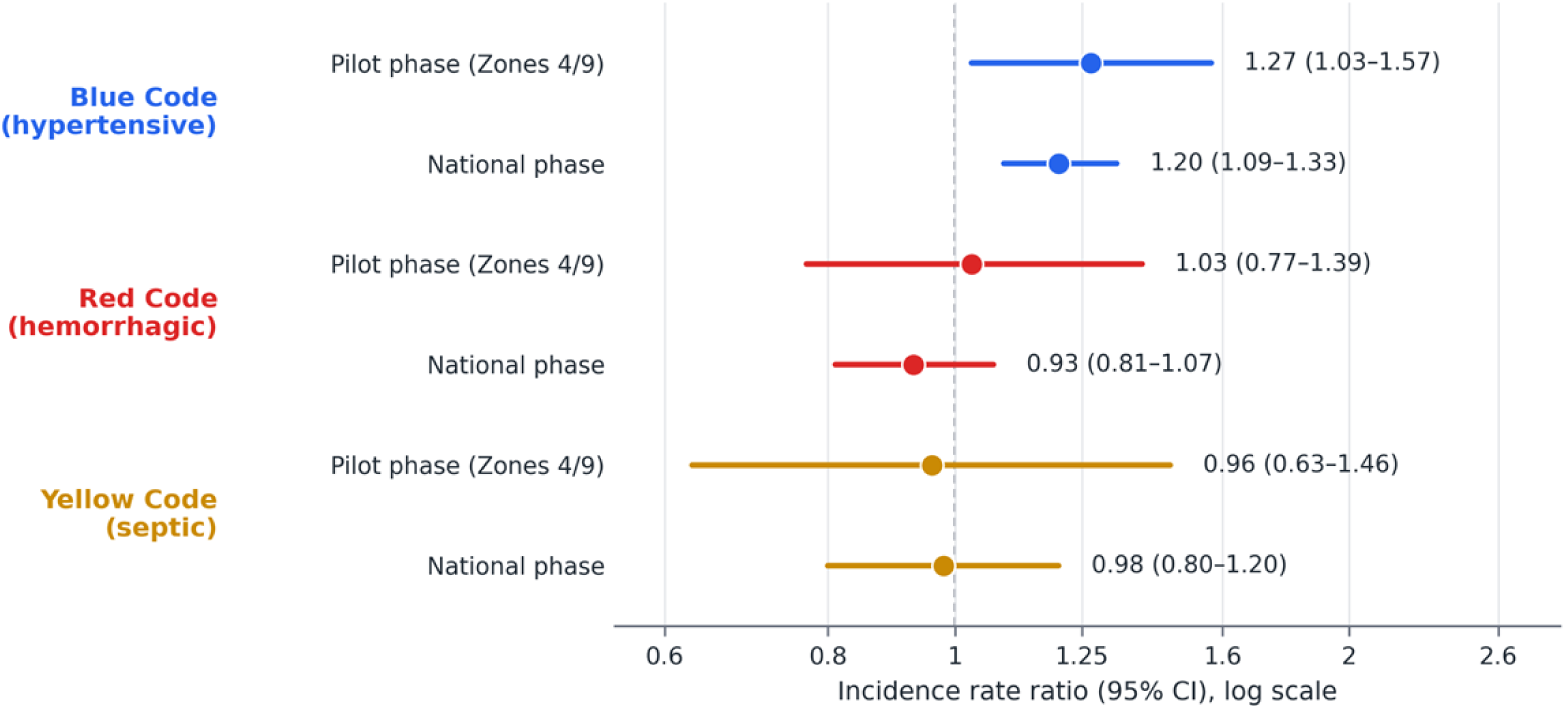
Incidence rate ratios (IRR) with 95% confidence intervals for obstetric complications, by intervention phase. Score MAMÁ, Ecuador, 2012–2024. Note. IRR: incidence rate ratio; CI: confidence interval; Z4/Z9: SENPLADES planning Zones 4 and 9 (pilot group). Dashed vertical reference line indicates the null value (IRR = 1). Blue Code: hypertensive disorders (ICD-10 O14–O15); Red Code: obstetric hemorrhage (O44–O46, O67, O72); Yellow Code: pregnancy-related sepsis (O85–O86, O41.1).

### Mortality analysis

Between 2012 and 2024, 817 maternal deaths were classified by obstetric code (Blue = 459; Red = 281; Yellow = 77) against the same national live-birth denominator used for the complications analysis (3 581 602 live births). Annual counts per code were small, particularly for Yellow Code (3–11 deaths/year), and the resulting rate series was correspondingly noisy: the combined rate across all three codes (Blue+Red+Yellow) fell from 35.7 per 100 000 live births in 2012 to 14.5 in 2016, rose again to 35.1 in 2020 — coincident with the COVID-19 pandemic — and fell to 13.9 by 2024.

Table 2 and Figure 2 present the incidence rate ratio for the national-level mortality change coincident with the 2016 rollout. None of the four models — Blue, Red, Yellow, or the pooled Blue+Red+Yellow measure — showed a statistically significant change, and all 95% CIs spanned the null value. For Red Code, the post-intervention slope was 22% per year (IRR 1.22; 95% CI 1.00–1.49; p = 0.051) — notable despite falling just short of significance, given the wide interval and small annual counts — consistent with a shift from a declining pre-intervention trend to a roughly flat trajectory after 2016, paralleling the null complications-rate finding for this code and Ministry surveillance data showing hemorrhage overtaking hypertensive disorders as Ecuador’s leading direct cause of maternal death by 2024 (15,16).

**Table 2.** Incidence rate ratio (IRR) for the association between the 2016 national rollout of Score MAMÁ and national maternal mortality.

| Code | IRR (2016) | 95% CI | p value |
| --- | --- | --- | --- |
| Blue (hypertensive) | 0.89 | 0.41 – 1.92 | 0.775 |
| Red (hemorrhagic) | 0.76 | 0.40 – 1.44 | 0.388 |
| Yellow (septic) | 1.62 | 0.56 – 5.32 | 0.396 |
| Pooled<br>(Blue+Red+Yellow) | 0.89 | 0.47 – 1.67 | 0.726 |

*Note. IRR: incidence rate ratio; CI: confidence interval. Estimates represent the level change in national annual maternal mortality coincident with the January 2016 national rollout of Score MAMÁ; the August 2015 pilot phase could not be isolated at this resolution. Blue Code: hypertensive disorders (ICD-10 O14–O15); Red Code: obstetric hemorrhage (O44–O46, O67, O72); Yellow Code: pregnancy-related sepsis (O85–O86, O41.1); Pooled (Blue+Red+Yellow): aggregate model summing cases across the three codes.*

**Figure 2.**
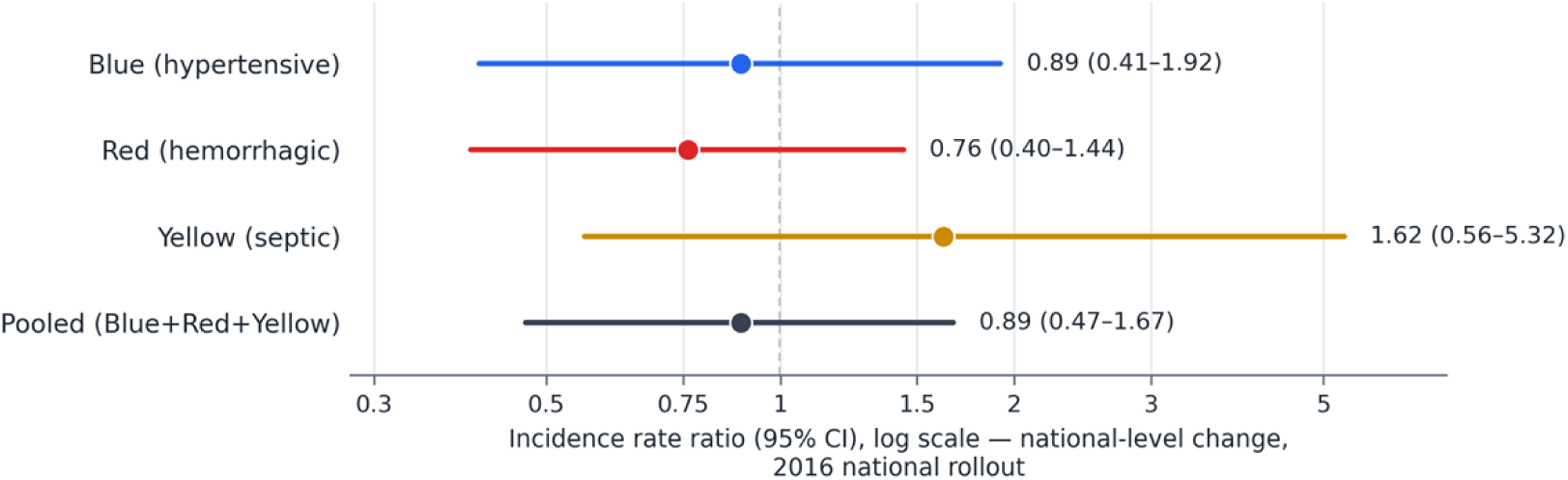
Incidence rate ratios (IRR) with 95% confidence intervals for national maternal mortality, associated with the 2016 national rollout of Score MAMÁ, by obstetric code. Note. IRR: incidence rate ratio; CI: confidence interval. Dashed vertical reference line indicates the null value (IRR = 1). Estimates represent the level change in national maternal mortality coincident with the January 2016 national rollout. Blue: hypertensive disorders (ICD-10 O14–O15); Red: obstetric hemorrhage (O44–O46, O67, O72); Yellow: pregnancy-related sepsis (O85–O86, O41.1).

Figure 3 displays the corresponding national annual maternal mortality trend by obstetric code across the full study period.

**Figure 3.**
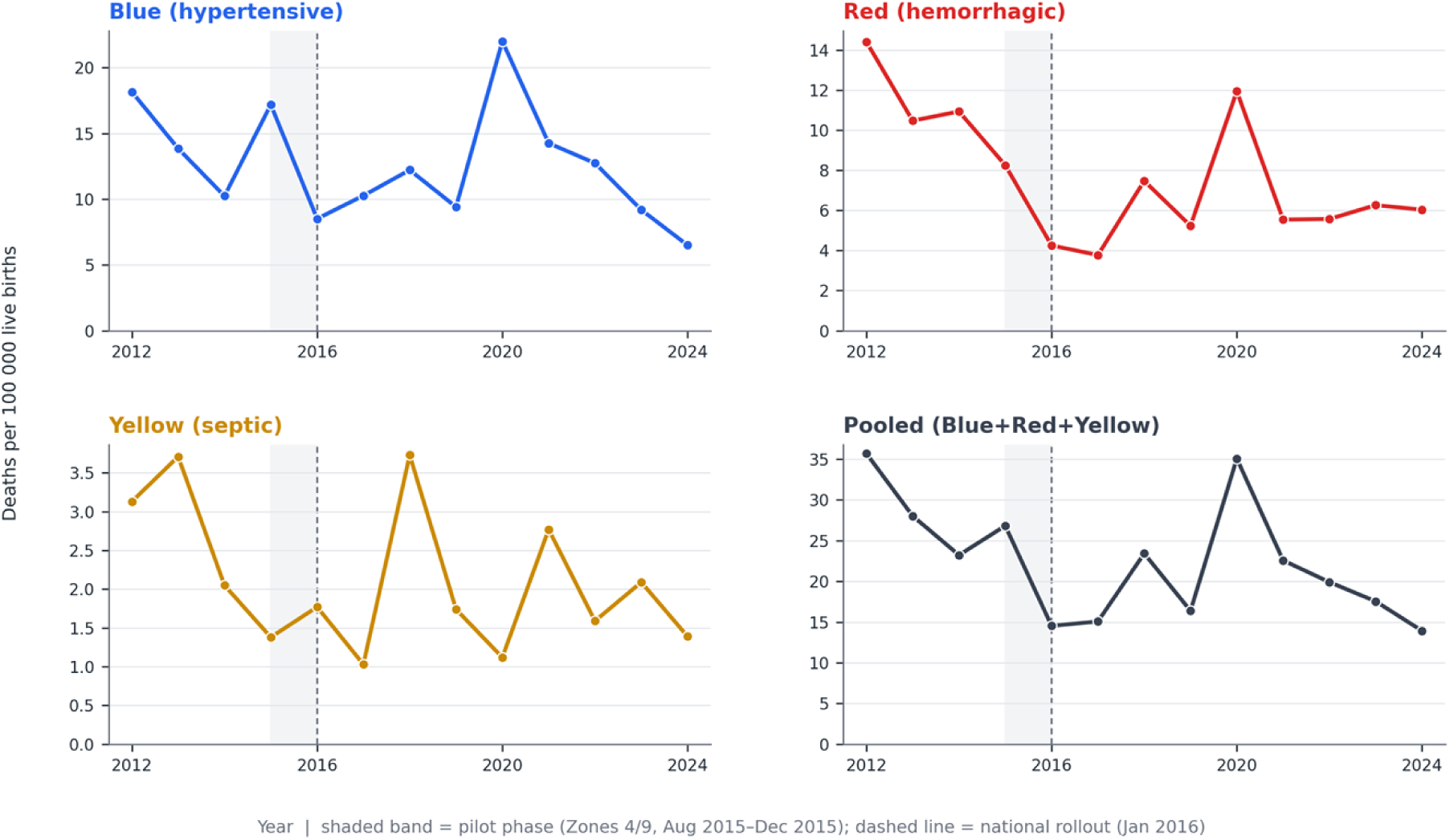
National maternal mortality rate per 100 000 live births by obstetric code. Note. Rates expressed as maternal deaths per 100 000 live births. Shaded band: pilot phase (Zones 4/9, Aug–Dec 2015); dashed line: national rollout (Jan 2016). Blue: hypertensive disorders (ICD-10 O14–O15); Red: obstetric hemorrhage (O44–O46, O67, O72); Yellow: pregnancy-related sepsis (O85–O86, O41.1).

### Sensitivity and robustness analyses

Five sensitivity analyses supported the robustness and specificity of the Blue Code finding (Methods). ACF/PACF diagnostics confirmed strong lag-1 residual autocorrelation (mean r = 0.33–0.54) decaying sharply thereafter (mean partial autocorrelation ≤0.21 at lag 2), supporting the AR1(1) structure. Restricted to the pre-intervention period, Blue Code showed a significant baseline trend difference between pilot and non-pilot zones (interaction p < 0.001); as this is already a covariate in the primary model, it does not bias the post-intervention IRR, though the trend-interaction term itself should not be over-interpreted (no such difference for Red or Yellow, p = 0.26 and p = 0.34). The seasonally adjusted variant left the Blue Code national effect essentially unchanged (IRR 1.26; p < 0.001) while attenuating the pilot-phase effect to borderline significance (IRR 1.22; p = 0.055); DHARMa tests showed adequate fit for Blue Code (p = 0.12) but persistent overdispersion for Red and Yellow even after seasonal adjustment (p < 0.001, p = 0.008); an alternative linear-dispersion specification for these two codes did not converge reliably. COVID-19 checks (pandemic-year indicator; exclusion of 2020–2021) left estimates essentially unchanged for all codes. A negative control outcome unrelated to Score MAMá’s protocols (diabetes complicating pregnancy, ICD-10 O24) showed no significant shift (IRR 0.94, p = 0.805; IRR 1.08, p = 0.492), supporting specificity of the Blue Code finding. On an absolute scale, the fitted national Blue Code rate exceeded the no-intervention counterfactual by 94.2 per 10 000 live births by December 2024 (Figure 4).

**Figure 4.**
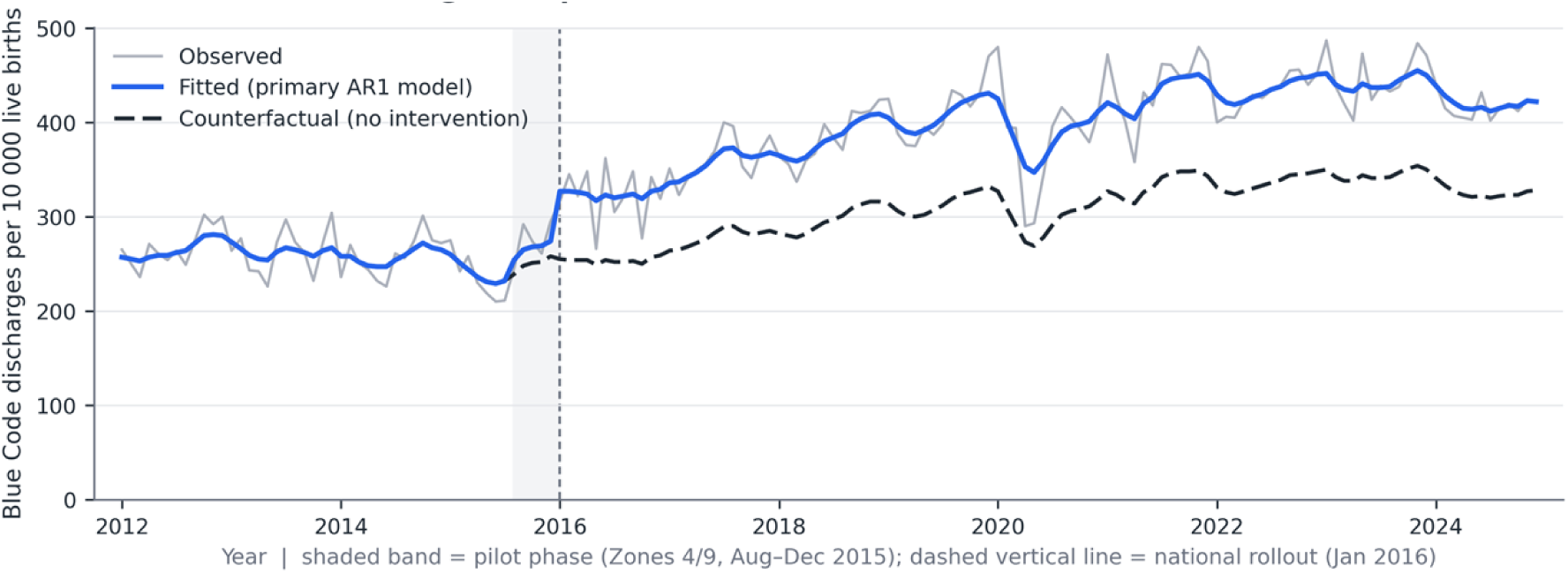
Blue Code (hypertensive disorders): national monthly observed, model-fitted, and no-intervention counterfactual discharge rate per 10 000 live births, 2012–2024. Note. Rates expressed as Blue Code (hypertensive disorders, ICD-10 O14–O15) hospital discharges per 10 000 live births, national monthly aggregate. Observed: raw monthly rate; Fitted: value predicted by the primary AR1 model; Counterfactual: value predicted by the same model with post_piloto and post_nacional fixed at 0 (i.e., the projected trajectory absent Score MAMÁ). Shaded band: pilot phase (Zones 4/9, Aug–Dec 2015); dashed line: national rollout (Jan 2016).

## Discussion

The central finding of this study is a clear dissociation between types of obstetric complication: implementation of Score MAMÁ was associated with a significant and sustained increase — in both the pilot and national phases — only in recorded Blue Code rates, while Red Code and Yellow Code remained unchanged. This differential pattern is key to correctly interpreting the nature of the observed effect.

A first hypothesis, implausible here, would be that Score MAMÁ caused a true deterioration in maternal health; this does not explain why the effect is confined to a single code, nor fits an intervention built on detection and clinical-response protocols. A second, more plausible hypothesis is increased ascertainment: a system standardizing routine blood-pressure measurement and proteinuria screening mechanically increases diagnosed preeclampsia-eclampsia cases, even without any change in underlying incidence, particularly at the milder end of the spectrum. We use “ascertainment” rather than “detection bias” deliberately: an increase in recorded cases following an intervention designed to improve clinical recognition is plausibly part of its intended mechanism rather than a methodological artifact, though our ecological design cannot distinguish between the two. This is consistent with prior evidence: early-warning systems perform well for hypertensive disorders in comparable Latin American settings (5), while their ability to identify sepsis has repeatedly been questioned (7), which could partly explain the null Yellow Code finding; severe hemorrhage, in turn, is acute and hemodynamically evident, rarely depending on a screening system to be recognized, which could partly explain the null Red Code finding. This mechanism may also operate administratively: a single, nationally mandated nomenclature likely also standardized how these events are coded in discharge and mortality records, a channel through which ascertainment can rise independently of true clinical incidence.

This finding suggests that the Blue Code rate indicator should not, on its own, be interpreted as a signal of deteriorating population-level maternal health following implementation of a surveillance system. It is also consistent with the growing regional prominence of hypertensive disorders as a cause of maternal mortality in Latin America (1,2): if Score MAMÁ improves early detection of preeclampsia, subsequent follow-up studies would be expected to show an associated reduction in case fatality from this code, even as the crude detection rate initially rises.

A question this evaluation should address directly, given that preventing maternal death is one of Score MAMÁ’s explicit objectives, is whether the observed change in complication rates was accompanied by any detectable change in maternal mortality. Our exploratory, national-level analysis found no statistically significant change in mortality for any code, nor in the pooled Blue+Red+Yellow measure, coincident with the 2016 rollout. This null result should not be read as evidence of no effect: at national-annual resolution, event counts were small (at most 59 deaths in a single year for any code), yielding wide confidence intervals and limited power to detect any but a very large effect — the same limitation, in more extreme form, that already qualifies the null complications-rate findings for Red and Yellow Code. One signal worth noting, though short of conventional significance, is a shift in Red Code mortality from a declining pre-2016 trend to a roughly flat trajectory afterward, directionally consistent with the absent complications-rate benefit for hemorrhage and with Ministry data showing hemorrhage overtaking hypertensive disorders as Ecuador’s leading direct cause of maternal death by 2024 (15,16). Taken together, Score MAMÁ’s demonstrated impact on ascertainment of hypertensive complications has not yet translated, or has not been shown to translate, into measurable national mortality reduction, with hemorrhage-related mortality warranting particular attention — timely given the Ministry’s 2025 protocol update (17).

## Limitations

This study has several limitations. First, as an ecological design using aggregated records, it cannot determine whether an additional Blue Code case reflects a genuinely new diagnosis or a previously unrecorded mild case, and no clinical outcome data are available to test the ascertainment hypothesis directly. Second, the numerator and denominator are not strictly commensurable, as detailed in Methods. Third, civil birth registration lags differentially affect the most recent years (2023–2024). Fourth, 0.10–0.14% of records lacked a valid cantonal code and were excluded. Fifth, Red and Yellow Code models retained residual overdispersion not fully resolved by the specifications explored, including seasonal adjustment; a real, smaller-magnitude effect being masked cannot be ruled out. Sixth, the study spans the COVID-19 pandemic (2020–2021); sensitivity checks (a pandemic-year indicator, and separately, excluding 2020–2021) left estimates materially unchanged for all codes, arguing against confounding, though subtler disruptions to care-seeking or coding cannot be fully excluded. Seventh, the complications analysis is limited to the zonal scale; cantonal-level and adolescent-pregnancy analyses are under development as complementary studies. Eighth, the mortality analysis is restricted to national-annual resolution, since canton-of-residence is available only for 2012–2014; it cannot isolate the pilot phase, evaluates only the 2016 breakpoint, and has limited power given the rarity of maternal death, so a null association should not be read as evidence of no effect (cause-of-death coding may also carry its own ascertainment variability, though agreement with INEC’s maternal-death flag was exact where comparable). Ninth, a significant pre-intervention trend difference between pilot and non-pilot zones was identified for Blue Code; though modeled as a covariate, residual confounding by unmeasured correlated zone-level factors cannot be fully excluded.

## Conclusions

Implementation of Score MAMÁ in Ecuador was associated with a selective, sustained increase in recorded rates of hypertensive complications, with no detectable change in hemorrhage or sepsis. A complementary, exploratory national-level analysis found no statistically significant change in maternal mortality for any code, though with limited statistical power given the rarity of maternal death. Taken together, this pattern is most consistent with increased ascertainment of preeclampsia-eclampsia, without yet-demonstrable evidence of a mortality benefit — a gap most notable for hemorrhage, which merits particular clinical and programmatic attention. Complementary studies incorporating clinical severity data, small-area spatial analysis, stratification by maternal age, and adequately powered rare-event methods for cause-specific mortality are needed to corroborate this interpretation.

## Declaration on the use of artificial intelligence

During preparation of this work, the authors used Claude (Anthropic) as a drafting assistant for data-analysis code and manuscript preparation, under the direct supervision and review of the corresponding and lead authors. All analytical decisions, interpretation of results, and the final scientific content are the responsibility of the named authors. Statistical analyses were performed in R 4.6.1 within RStudio 2026.08.1.

## Data availability statement

The aggregated monthly panel dataset (zone- and obstetric-code-level complication counts, live-birth denominators, and the national annual maternal mortality series by code) underlying the findings of this study is deposited in Zenodo and available at https://doi.org/10.5281/zenodo.23018105. The original source microdata — Live Birth Statistics, Hospital Discharge records, and General Mortality Statistics — are publicly available from Ecuador’s National Institute of Statistics and Census (INEC) at https://www.ecuadorencifras.gob.ec/institucional/home/, subject to INEC’s terms of use. Analysis code used to generate the results of this article is available from the corresponding author upon request.

## Author contributions

Conceptualization: A.S.H.A., A.W.G.V. Methodology: A.S.H.A., A.W.G.V. Software: A.S.H.A. Formal analysis: A.S.H.A. Investigation: A.W.G.V. Data curation: A.S.H.A., E.S.L.M. Writing – original draft: A.S.H.A., E.S.L.M. Writing – review & editing: M.A.G.S. Supervision: L.G.P. Resources: L.G.P. Validation: M.A.G.S. Project administration: A.A.T.S. All authors read and approved the final manuscript and meet the ICMJE criteria for authorship, as attested in the signed RPSP/PAJPH Author Declaration Form retained by the corresponding author.

## Funding

This research received no specific grant from any funding agency in the public, commercial, or not-for-profit sectors. The funders (not applicable) had no role in study design, data collection and analysis, decision to publish, or preparation of the manuscript.

## Competing interests

The authors have declared that no competing interests exist.

## Notes

### Competing Interest Statement

The authors have declared no competing interest.

### Author Declarations

The study protocol was reviewed by the Research Ethics Committee on Human Subjects (CEISH, Comité de Ética de Investigación en Seres Humanos) of the International University of Ecuador (UIDE). The committee determined that this study, which used exclusively secondary, aggregated, anonymized administrative data with no individual patient identification or direct patient contact, was exempt from full committee review.

